# Personalizing renal replacement therapy initiation in the intensive care unit: a reinforcement learning-based strategy with external validation on the AKIKI randomized controlled trials

**DOI:** 10.1101/2023.06.13.23291349

**Authors:** François Grolleau, François Petit, Stéphane Gaudry, Élise Diard, Jean-Pierre Quenot, Didier Dreyfuss, Viet-Thi Tran, Raphaël Porcher

## Abstract

**Background:** Trials sequentially randomizing patients each day have never been conducted for renal replacement therapy (RRT) initiation. We used clinical data from routine care and trials to learn and validate optimal dynamic strategies for RRT initiation in the intensive care unit (ICU).

**Methods:** We included participants from the MIMIC-III database for development, and AKIKI and AKIKI2 (two randomized controlled trials on RRT timing) for validation. Participants were eligible if they were adult ICU patients with severe acute kidney injury, receiving invasive mechanical ventilation, catecholamine infusion, or both. We used doubly-robust estimators to learn when to start RRT after the occurrence of severe acute kidney injury given a patient’s evolving characteristics—for three days in a row. The ‘crude strategy’ aimed to maximize hospital-free days at day 60 (HFD60). The ‘stringent strategy’ recommended initiating RRT only when there was evidence at the 0.05 threshold that a patient would benefit from initiation. For external validation, we evaluated the causal effects of implementing our learned strategies *versus* following current best practices on HFD60.

**Results:** We included 3 748 patients in the development set (median age 69y [IQR 57– 79], median SOFA score 9 [IQR 6–12], 1 695 [45.2%] female), and 1 068 in the validation set (median age 67y [IQR 58–75], median SOFA score 11 [IQR 9–13], 344 [32.2%] female). Through external validation, we found that compared to current best practices, the crude and stringent strategies improved average HFD60 by 13.7 [95% CI-5.3–35.7], and 14.9 [95% CI - 3.2–39.2] days respectively. Contrasted to current best practices where 38% of patients initiated RRT within three days, with the stringent strategy, we estimated that only 14% of patients would.

**Conclusion:** We developed a practical and interpretable dynamic decision support system for RRT initiation in the ICU. Its implementation could improve the average number of days that ICU patients spend alive and outside the hospital.

## Introduction

In intensive care units (ICU), acute kidney injury (AKI) affects about one in two patients, and its onset is associated with high mortality and long-term sequelae (1). Renal replacement therapy (RRT) is an invasive but potentially life-saving treatment for AKI (2). Because AKI is a heterogeneous and rapidly evolving syndrome (3), controversies on the timing and selection of patients for initiating RRT have long prevailed (4). In the last decade, multicenter randomized trials, compared early versus delayed RRT initiation strategies, but the analyses (5–7) and meta-analyzes (8, 9) of these trials failed to show significant differences in patient-important outcomes at the population level. As such negative trial findings are widespread in critical care, identifying individualized treatment effects has been judged a research priority (10).

Physicians’ attempts to deliver timely interventions tailored to patients’ characteristics have a long history (11). While in some diseases, biological insight proved decisive in moving precision medicine forward (12), AKI—due to its heterogeneous syndromic nature—is less amenable to this approach. Recently, authors proposed algorithms for RRT initiation in the ICU (13, 14), but the need for validated data-driven decision support tools remains (15). Previously, we developed a decision support tool based on clinical trial data and considered the static case where the decision to initiate RRT is only pondered at AKI onset (16). Yet, for such decision tools to be actionable and consistent with practice, they must go beyond the static case and account for the fundamentally dynamic nature of AKI. In fact, when a decision support tool recommends not initiating RRT for a given patient on a given day, it ought to re-evaluate its recommendation on the next day considering the evolution of the patient’s characteristics.

To learn an optimal RRT initiation strategy under this setting, the ideal method would be to conduct a Sequential Multiple Assignment Randomized Trial (SMART) where AKI patients are sequentially randomized each day to either initiate treatment or not (17). Due to cost, time, and practical constraints, SMART trials have never been conducted in the ICU. However, recent developments in statistics and computer science provided robust methods to learn and evaluate optimal treatment initiation strategies from observational data (18–20). To our knowledge, only a single monocenter study has analyzed clinical data in an attempt to develop a dynamic decision support system for RRT initiation (21).

In this paper, we used reinforcement learning methods on data from electronic health records to estimate optimal dynamic strategies for RRT initiation in ICU patients with AKI. Then, in an external validation step, we used data from two large multicenter randomized trials on RRT timing to estimate the benefit of implementing these strategies.

## Methods

### Sources of data

The development sample included participants from the Multi-Parameter Intelligent Monitoring in Intensive Care III (MIMIC-III) database. MIMIC-III is a project maintained by the Laboratory for Computational Physiology at the Massachusetts Institute of Technology which contains routinely collected data from 61,051 distinct ICU admissions of adult patients admitted between 2001 and 2012 (22). For reproducibility, we used the database’s official code repository to extract all relevant variables (23). As out-of-hospital mortality was not available in the latest version of the MIMIC project, we used MIMIC-III version 1.4.

The validation sample included participants from the AKIKI and AKIKI2 trials, two multicenter RCTs conducted in France (5, 24). The AKIKI trial was conducted at 31 ICUs from Sept 2013 through Jan 2016 and recruited 619 patients with stage 3 KDIGO-AKI who required mechanical ventilation, catecholamine infusion, or both. Included patients were 1:1 randomized to either an early RRT initiation strategy or to a standard-delayed initiation strategy. The AKIKI2 trial was embedded in a cohort recruiting at 39 ICUs from May 2018 through Oct 2019. In AKIKI2, eligibility criteria for the cohort were identical to the eligibility criteria from the original AKIKI trial. Of the 767 patients included in the cohort, 278 met one or more randomization criteria (oliguria for more than 72h or blood urea nitrogen concentration greater than 112 mg/dL) and were 1:1 allocated to either a standard-delayed RRT initiation strategy or to a more-delayed strategy.

### Population

Eligible patients were adults (18 years of age or older) hospitalized in the ICU with stage 3 KDIGO AKI who were receiving (or had received for this episode) invasive mechanical ventilation, catecholamine infusion, or both. Staging in the KDIGO classification was based on serum creatinine and/or urine output with higher stages indicating greater severity (25). As the latest clinical guidelines recommend a standard-delayed strategy of RRT initiation (26), we chose this strategy as the reference “best practice” upon which to improve. Precisely, our target population was made of individuals whose physicians implemented a standard-delayed strategy. In both AKIKI trials, the standard-delayed strategy suggested initiating RRT if one of the following criteria occurred: severe hyperkalemia and/or metabolic acidosis, pulmonary oedema resistant to diuretics, oliguria for more than 72 hours, blood urea nitrogen level higher than 112 mg per deciliter. In the current study, we used the same exclusion criteria as in the AKIKI trials i.e., moribund state (patient likely to die within 72h), end-stage kidney disease (i.e., patient with creatinine clearance < 15ml/ml), patients having received RRT before inclusion, and patients already included at a previous date.

### Setup and timepoints for learning dynamic RRT initiation strategies

From a clinical standpoint, the decision to start RRT is considered difficult in the first days following severe AKI. After three days, this decision often becomes straightforward, as most patients have either recovered or deteriorated. We focused on developing a when-to-treat strategy for RRT initiation in the first 72 hours following the onset of severe AKI (i.e., stage 3 KDIGO-AKI). Specifically, we learned a strategy that—for three days in a row after severe AKI onset—assessed the need to start RRT given a patient’s evolving characteristics. Our strategy was non-stationary i.e., the decision rules for RRT initiation could differ depending on the day. We considered three decision timepoints at 0, 24, and 48 hours after severe AKI onset (**Figure 1**). The strategy was developed so that, at each timepoint, it used clinical and biological information gathered prior to this timepoint as inputs and outputted a recommendation to either initiate RRT within 24 hours or not. We considered that once RRT had been recommended (or initiated in contradiction with the strategy’s recommendation), the strategy would persist in recommending RRT for all subsequent decision timepoints. This so-called regularity in the strategy’s behavior indicates that we did not consider when to stop RRT in the three days following severe AKI onset.

**Figure 1.**
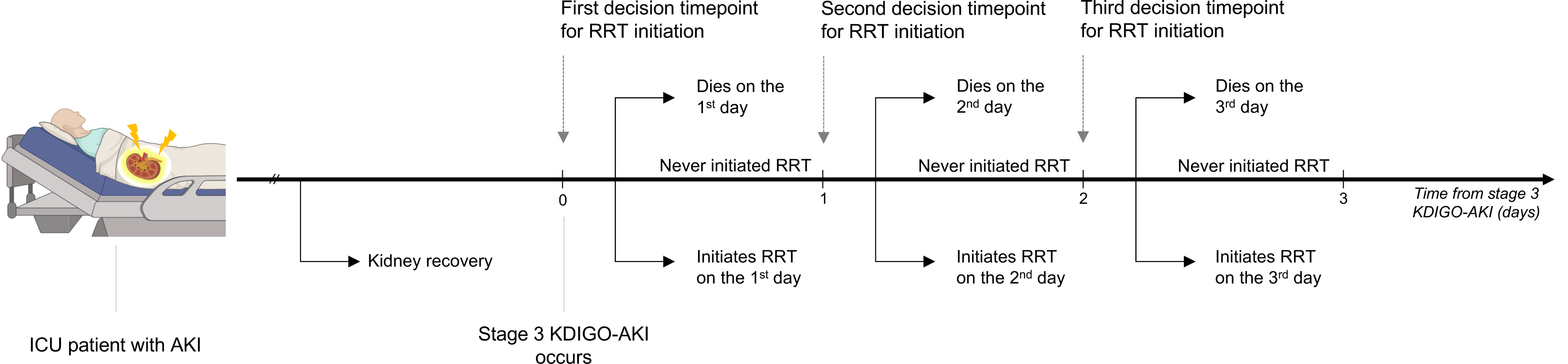
Possible trajectories of a single patient with acute kidney injury in our learning setup. The first decision timepoint is defined as the time when stage 3 KDIGO-AKI occurs. In our setup, for a patient with stage 3 KDIGO-AKI, the decision rule to initiate RRT mimics that of clinicians i.e., decisions are re-evaluated every day—for three days in a row–given patients’ evolving characteristics. Note that at a given decision timepoint a decision needs to be made only if a patient has neither initiated RRT nor died earlier. AKI=Acute Kidney Injury. ICU=Intensive Care Unit. KDIGO=Kidney Disease Improving Global Outcomes. RRT=Renal Replacement Therapy.

### Primary outcome

The primary outcome was hospital-free days at day 60 (HFD60). This outcome was chosen because i) it was a good compromise between patient-centeredness and pragmatism (27); and it reduced the risk that the learned strategy had unexpected side effects—a well-known issue in reinforcement learning (28). For instance, using short-term mortality as a primary outcome, the model may learn a strategy that maximizes survival at the cost of keeping patients alive in the ICU as long as possible.

### Learning an optimal strategy

To learn an optimal strategy, we used a doubly robust estimator with weighted least squares (dWOLS) (29). This method relies on estimating blip functions for each decision timepoint. Given the evolving characteristics of an individual up to timepoint *t*, a blip function predicts the effect of initiating RRT at *t* versus not initiating it at *t* but taking optimal treatment decisions from timepoint *t* + 1 onwards. We derived two strategies from the estimated blip functions. We termed “crude” the strategy that recommended RRT initiation to the patients with positive values of blips, and “stringent” the strategy that recommended initiating RRT only when there was evidence at the 0.05 significance level that a patient would benefit from RRT initiation (i.e., positive lower bound for the blip’s 95% confidence interval). As stated before, once RRT initiation was recommended, both strategies persisted in recommending RRT regardless of the blips at subsequent timepoints. Patients who died within three days of AKI were excluded from the development sample, considering no relevant information would be learned from these patients’ data. Indeed, it seemed unlikely that patients who died within three days of severe AKI could have been discharged from the hospital under a different RRT initiation strategy: we expected their outcome to be the same under all strategies (HFD60 is zero for all patients who die in the hospital). However, these patients were not excluded from the validation sample, to avoid time-dependent selection bias. More details on dWOLS estimation and inference are given in the appendix (pp 3-4).

### External validation

To match our target “best practice” population, we included all patients from AKIKI and AKIKI2 who had received a standard-delayed strategy. As the AKIKI2 patients randomized to a more-delayed strategy were compatible with a standard-delayed strategy until they met a randomization criterion, we excluded these patients but duplicated the patients randomized to the standard-strategy arm according to the cloning and censoring principle used for emulating target trials from observational data (30). To estimate hospital mortality and the proportion of patients who would initiate RRT within three days under a strategy, we used importance sampling for policy evaluation in reinforcement learning (31).

To evaluate the effect of new strategies on HFD60, we considered current best practices (i.e., the standard-delayed strategy from the AKIKI trials) as a common control and compared it to the following three strategies: i) the crude strategy, ii) the stringent strategy, and iii) a strategy that recommends initiating RRT in all patients within 24 hours after severe AKI onset. We estimated the causal effect of implementing each of these strategies compared to current best practices using the cross-fitted advantage doubly robust estimator for strategy evaluation with terminal states (19). This estimator allows estimating the mean difference in the outcome that would have been observed under any given strategy and the outcome observed under a reference strategy. We provide more details on importance sampling for policy evaluation and the advantage doubly robust estimator in the appendix (pp 4-5).

### Ethical approval and research transparency

The MIMIC-III analysis received approval from the Institutional Review Boards of the Massachusetts Institute of Technology and Beth Israel Deaconess Medical Center (BIDMC). The AKIKI and AKIKI2 trials received approval from competent French legal authority (Comité de Protection des Personnes d’Ile de France VI, ID RCB 2013-A00765-40, NCT01932190 for AKIKI and ID RCB 2017-A02382-51, NCT03396757 for AKIKI 2) and participants provided written informed consent to take part in the study. The funding sources were not involved in the study design; collection, analysis, and interpretation of data; writing of the manuscript; or the process of submission for publication. Two authors (FG, RP) had full access to all the data in the study and take responsibility for the integrity of the data and the accuracy of the analysis. Analyses were conducted using R version 4.2.1 for strategy learning as well as plotting, and Python 3.8.8 for strategy evaluation. The code used in this study is available at https://github.com/fcgrolleau/dynamic-rrt.

## Results

### Learning optimal dynamic strategies for RRT initiation

#### 1. Patients

From 2001 through 2012, a total of 3 748 ICU patients with AKI recruited at a tertiary teaching hospital (BIDMC — Harvard Medical School) met eligibility criteria and were included in the development set (**Figure S1, Panel A**). Almost half of individuals were females (n=1 695; 45.2%). At enrollment, patients had a mean SOFA score of 9 (interquartile range [IQR], 6–12). All patients had severe AKI (i.e., stage 3 KDIGO-AKI) which diagnosis was most often based on urine output (n=3 328; 88.8%). At enrollment, the median serum creatinine and urine output were 1.40 mg/dL (IQR, 0.90–2.40) and 0.28 ml/kg/h (IQR, 0.22–0.29) respectively. During the follow-up, 400 (10.7%) patients initiated RRT within three days of severe AKI, and 892 (23.8%) died during hospitalization. The mean and median HDF60 were 33.6 and 42.9 days, (IQR, 0.9–51.7) respectively. Additional baseline and evolving characteristics for these patients are given in **Table 1**.

**Table 1.**
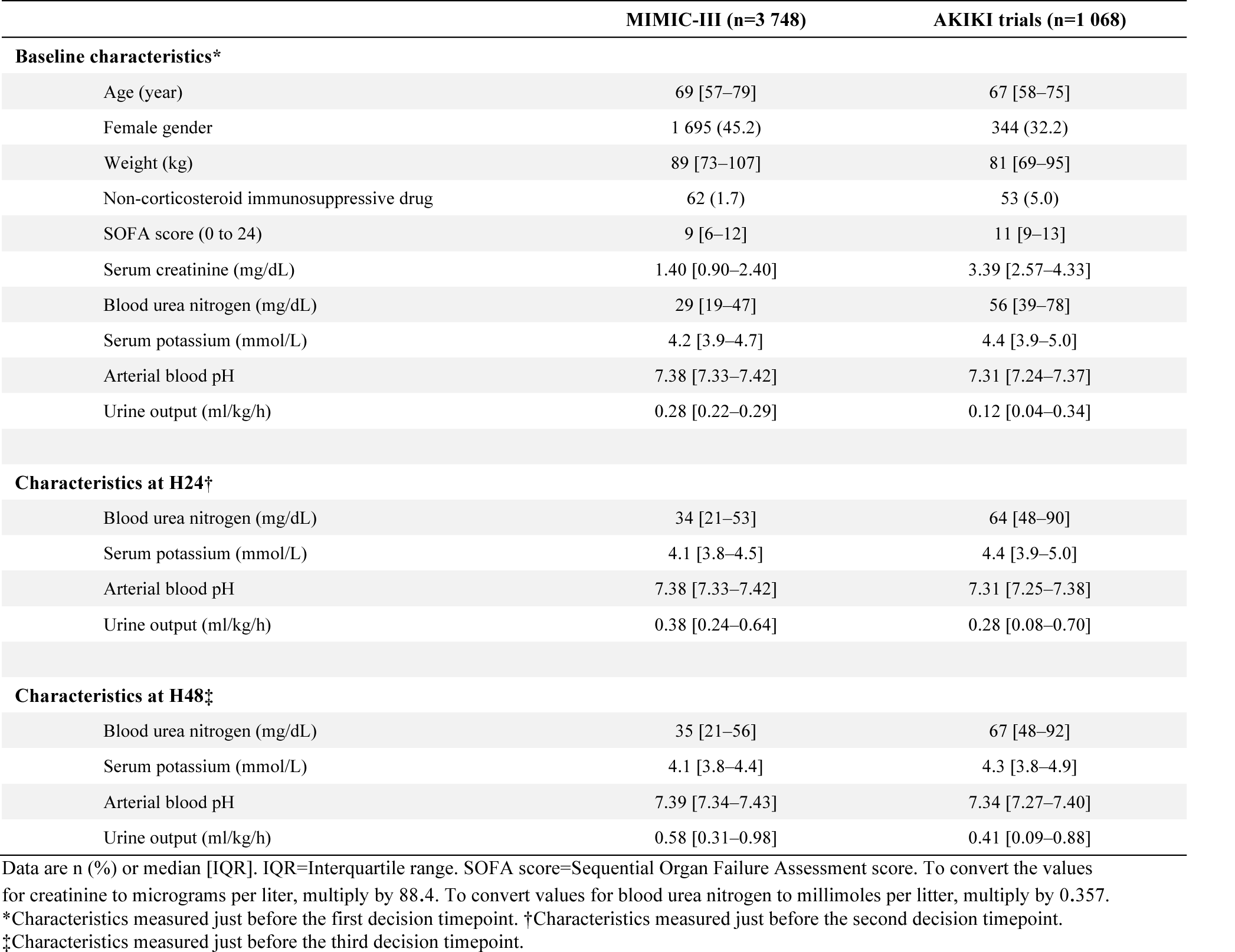
Baseline and evolving characteristics of the patients from the development set (MIMIC-III) and the validation set (AKIKI trials).

#### 2. Learned strategies

For patients with severe AKI who have never initiated RRT at a given decision timepoint, decision rules whether to initiate RRT in the next 24 hours were derived from the models we estimated at each timepoint (**Figure 1**). Estimated parameters of the so-called blip functions are given with didactical instructions for calculations in **Table 2** (their covariances are given in **Table S1**). In **Figure 2A** we display the recommendations from two learned strategies (i.e., our crude and stringent strategies) along with the uncertainty in the recommendation for each patient in the development set. We present in **Table 3**, three illustrative examples where the learned strategies were applied for individualizing the decision to initiate RRT within 72 hours of severe AKI. The apparent effect (i.e., in the development set) of implementing our crude strategy versus implementing the MIMIC-III RRT initiation strategy was a 6.6 days improvement in mean HFD60.

**Figure 2.**
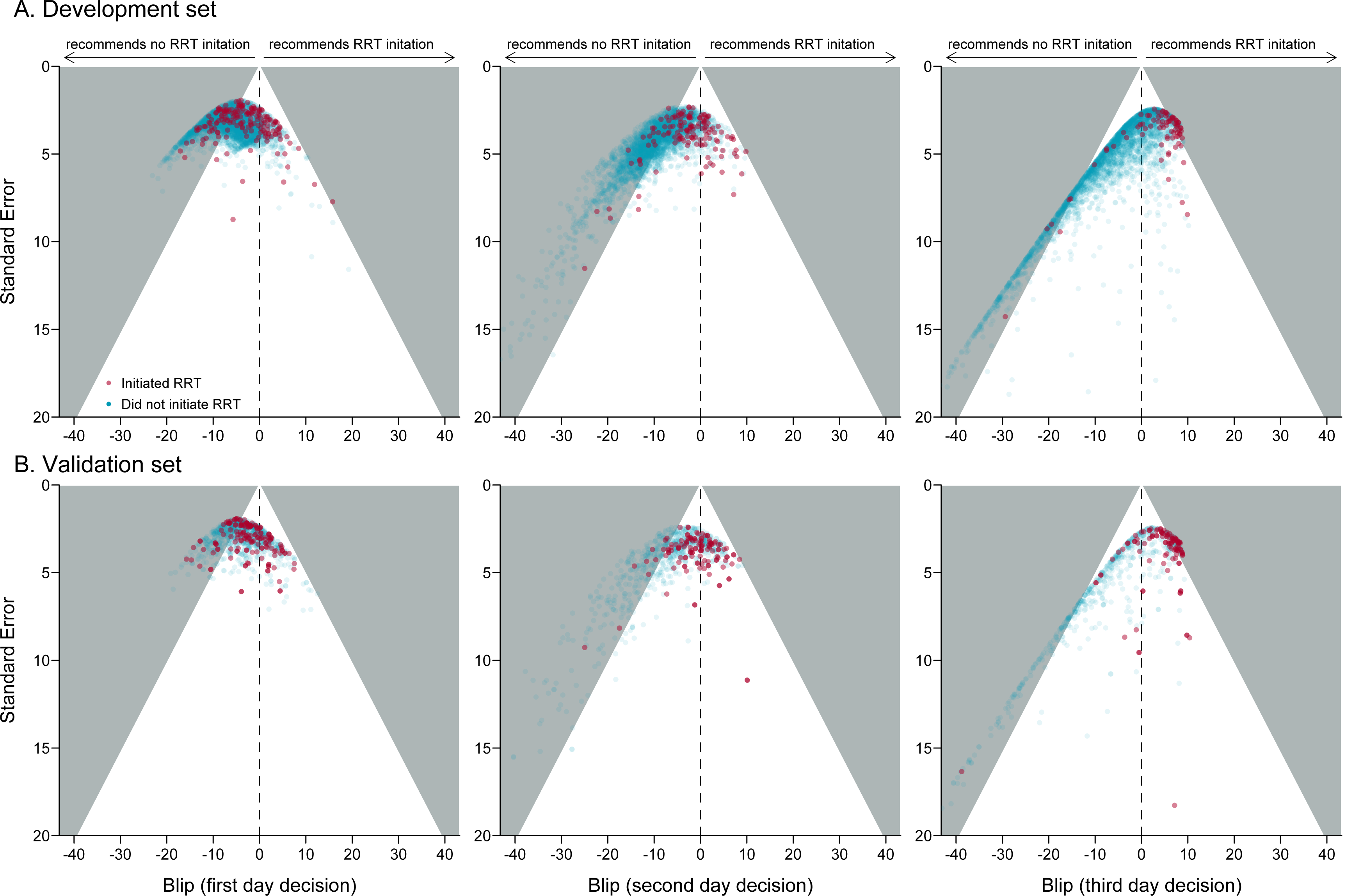
Recommendations from the learned strategies for patients in the development set (Panel A) and in the validation set (Panel B). Each dot corresponds to a patient for whom a decision whether to initiate RRT needed to be made at the first (left-hand panels), second (middle panels), or third (right-hand panels) decision timepoint. Dot colors depict the RRT prescription observed for these patients. On the on *x*-axis, predicted blips indicate on a HFD60 scale the magnitude of individual-patient harm (negative blips) or benefit (positive blips) from initiating RRT at a particular timepoint. Vertical dashed lines indicate no effect. Uncertainty in the individual-patient blips is represented on *y*-axis. Dots falling in grey-shaded aeras represent patients for whom there is evidence of either harm (left-hand aeras), or benefit (right-hand aeras) from RRT initiation at the 0.05 alpha level. The crude strategy would recommend initiating RRT at a given timepoint if a patient’s dot fell on the right-hand side of the dashed line. On the other hand, the stringent strategy would recommend initiating RRT at a given timepoint only if a patient’s dot fell in the right-hand grey-shaded aera.

**Table 2.**
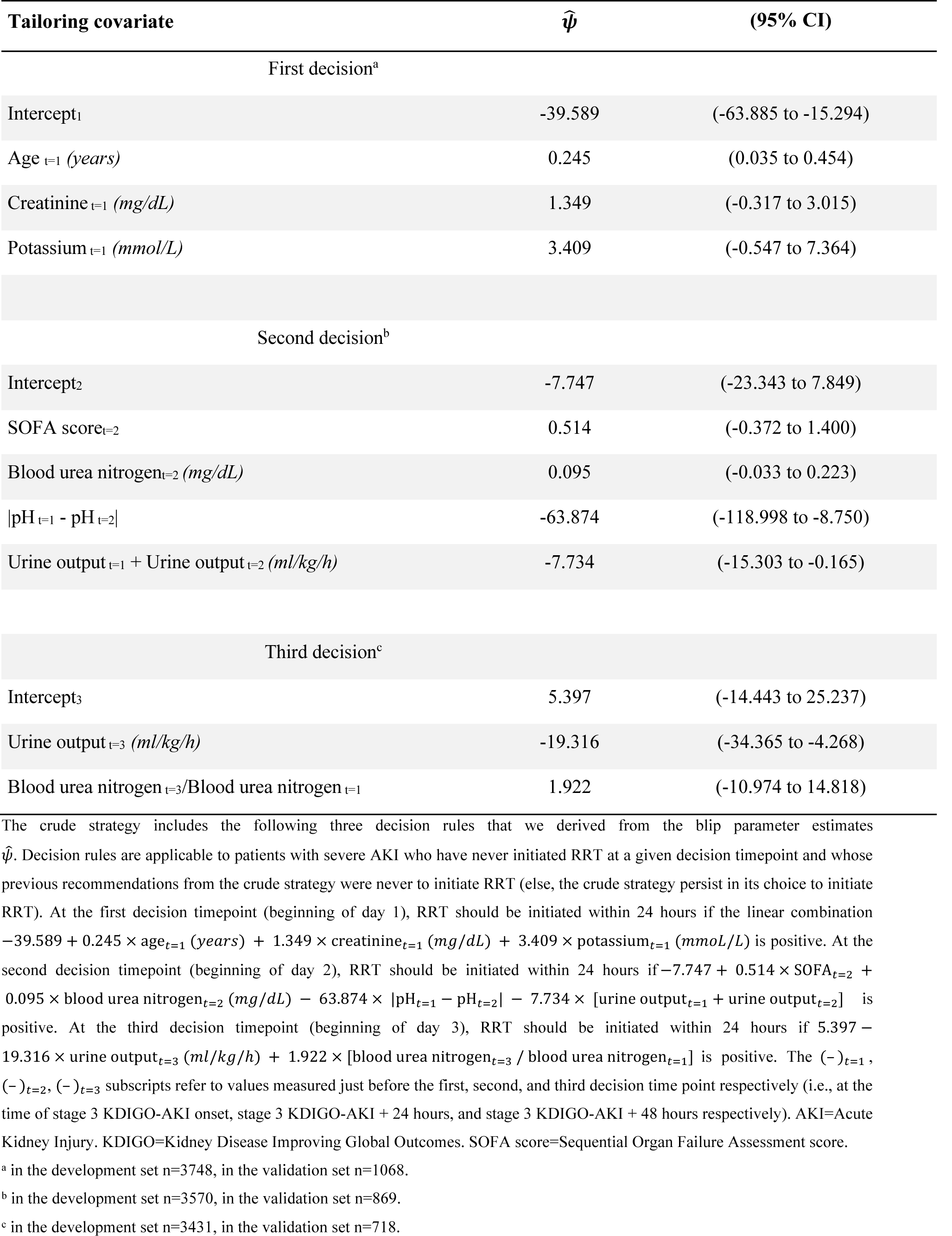
Blip parameter estimates from the learned strategies. Estimations based on the multiple imputation analysis of one hundred data sets.

**Table 3.**
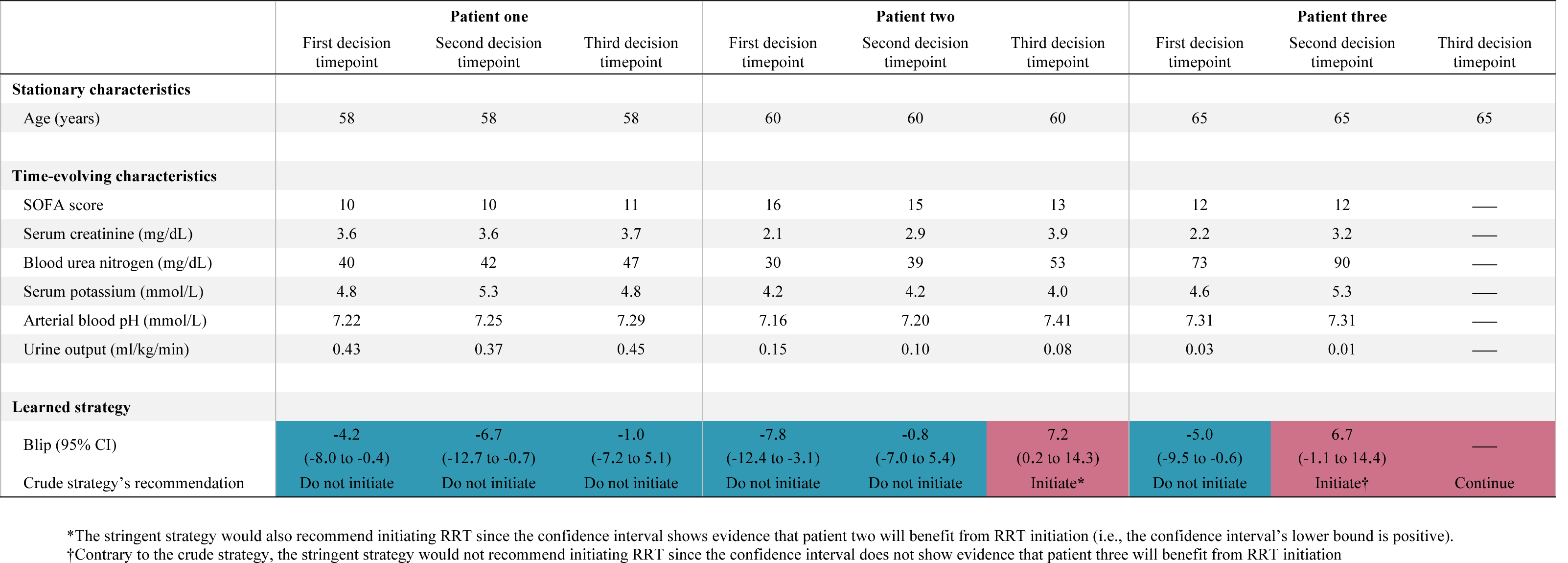
Use of the learned strategies for individualized decision-making in three illustrative examples. In patient one (a man aged 58 years), disease severity (as described by SOFA score and arterial blood pH) and kidney function (as described by blood urea nitrogen, serum creatinine, and urine output) remain stable, and the crude strategy suggests against initiating RRT in the 72 hours following stage 3 KDIGO-AKI onset. In patient two (a woman aged 60 years), disease severity lessens over time, but kidney function deteriorates, and the crude strategy suggests initiating RRT on the third day following stage 3 KDIGO-AKI. In patient three (a woman aged 65 years), disease severity is stabilized 24 hours after stage 3 KDIGO-AKI onset, but kidney function has become critical, and the crude strategy suggests initiating RRT on the second day. Note that once a learned strategy recommends initiating RRT it persists in its recommendation until the third day regardless of patients’ subsequent characteristics. AKI=Acute Kidney Injury. KDIGO=Kidney Disease Improving Global Outcomes. RRT=Renal Replacement Therapy. SOFA score=Sequential Organ Failure Assessment score.

### External validation

#### 1. Patients

From 2013 through 2019, a total of 931 unique ICU patients with AKI from the AKIKI and AKIKI2 trials met our predefined eligibility criteria and were included in the validation set. After cloning and censoring, these corresponded to a sample of 1 068 individuals from a population who have received current best practices (i.e., a standard-delayed strategy, *see* **Figure S1, Panel B**). About a third of individuals (n=344; 32.2%) from the validation set were females. For most patients, severe AKI was associated with septic shock and the mean SOFA score was 11 (IQR, 9–13). A drop in urine output triggered stage 3 KDIGO-AKI diagnosis in 401 patients (37.5%). At enrolment, the median serum creatinine and urine output were 3.39 mg/dL (IQR, 2.57–4.33) and 0.12 ml/kg/h (IQR, 0.04–0.34) respectively. During follow-up, 482 (45.1%) died during hospitalization, while 405 (38%) and 99 (20.5%) respectively initiated RRT or died within three days of severe AKI. The mean HFD60 was 14.2 days (median 0, IQR, 0–30.3).

#### 2. External validation of the learned strategies

In the external validation population, we estimated that, under our crude strategy, 41% of patients would die during hospitalization and 53% would initiate RRT within three days. Under our stringent strategy, we estimated that 38% of patients would die during hospitalization and 14% of patients would initiate RRT within three days. Recommendations from the learned strategies along with the uncertainty in individual-patient recommendations are given for all patients in the validation set in **Figure 2B**. The discrepancies between current best practices and the recommendations from the learned strategies are shown in **Figure 3**. We found that compared to current best practices (i.e., the standard-delayed strategy from the AKIKI trials), our crude and stringent strategies yielded a 13.7 days and 14.9 days improvement in mean HFD60 respectively (**Figure 4**).

**Figure 3.**
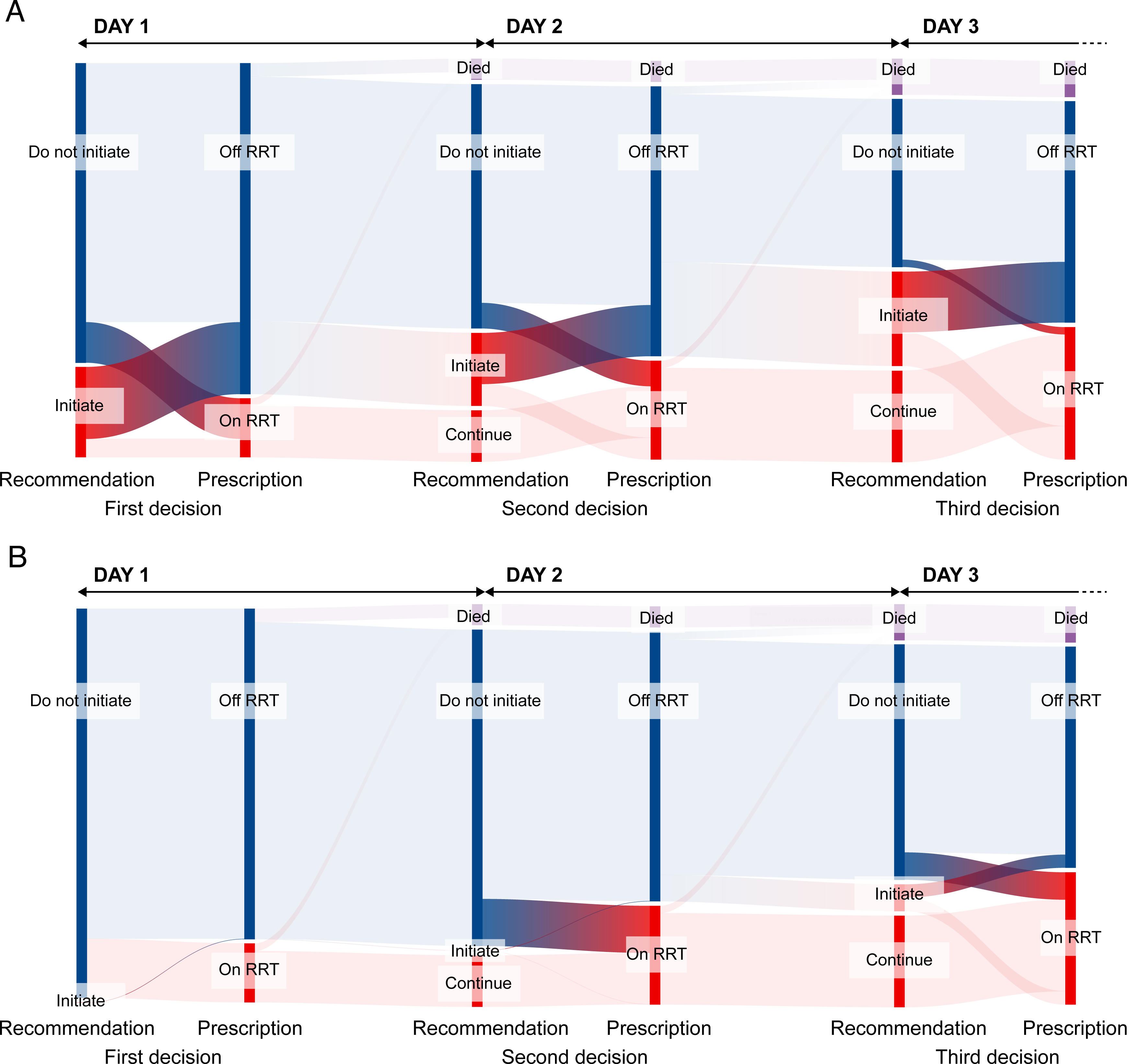
Comparison of recommendations from the crude (Panel A) or stringent (Panel B) strategy and the prescriptions received in the validation set. Prescriptions received are denoted ‘On RRT’ or ‘Off RRT.’ The bar heights represent the proportions of patients in each category. At each decision timepoint, recommendation and prescription of RRT appear in red while the absence of recommendation or prescription of RRT is shown in blue. Discrepancies between recommendations and prescriptions are shown in brighter colors. Note that when patients initiated RRT (sometimes in contradiction with the strategy’s recommendation) the strategy never recommends stopping it afterward.

**Figure 4.**
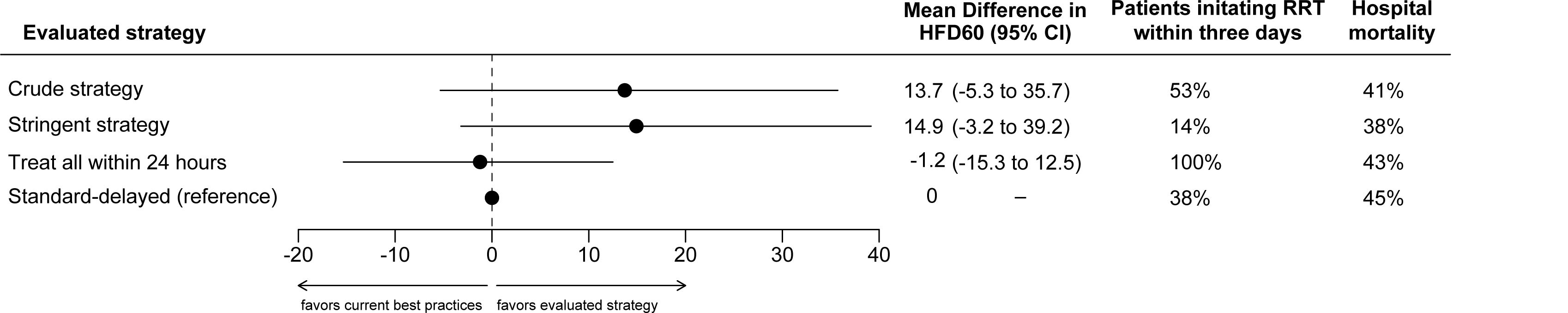
External validation of the learned strategies’ benefit as compared to current best practices (i.e., a standard-delayed strategy). The mean difference in HFD60 represent the causal effects of implementing a strategy compared to current best practices. The “crude strategy” refers to the strategy derived from the blip parameter estimates given in Table 2. The “stringent strategy” refers to a strategy that recommends initiating RRT only when there is evidence at the 0.05 threshold that a patient will benefit from RRT initiation. The “treat all within 24 hours strategy” designates a strategy to initiate RRT in all patients within 24 hours regardless of emergency criteria. HFD60=Hospital-Free Days at day 60. CI=Confidence Interval. RRT=Renal Replacement Therapy.

## Discussion

### Summary of findings

In this study, we used electronic health record data to learn dynamic RRT initiation strategies for ICU patients with severe AKI. Then, using data from two large RCTs of RRT timing we conducted external validation: compared to current best practices (i.e., a standard-delayed strategy), we found that the crude strategy may improve HFD60 by 13.7 days on average. Note that even though the crude strategy may recommend RRT initiation sooner than the standard-delayed strategy, it is not an early strategy. Consistent with previous trials (5–7), we showed that a strategy that recommends RRT initiation in all patients within 24 hours of severe AKI may yield outcomes similar to or worse than that of a standard-delayed strategy. In contrast to early strategies, the crude strategy identified that only 53% of patients required RRT initiation in the three days following severe AKI. Of note, in the STARRT-AKI and AKIKI arms corresponding to current best practices (i.e., the arms termed standard and delayed respectively), rates of RRT initiations a week after severe AKI were 59% and 55%. We believe that the benefit of the crude strategy stems from its ability to identify earlier the patients who will ultimately require RRT. That said, we found that the stringent strategy may also improve patients’ HFD60 all the while reducing RRT prescriptions in the three days following severe AKI. This suggests that the individual-patient confidence intervals given by the crude strategy provide important information for deciding the initiation of RRT. Entailing less frequent usage of RRT, the stringent strategy could have the benefit of not only improving patient-important outcomes but also saving health resources.

Our methodology aimed at developing interpretable linear decision rules together with confidence intervals to guide clinicians at the bedside. For greater transparency and interpretability, we released a user-friendly online implementation of our learned strategies at http://dynamic-rrt.eu. Using the time-varying characteristics of a patient as input, clinicians can with this web application obtain individual-patient recommendations from the crude strategy along its 95% confidence intervals. With respect to interpretability, we noticed that on the first day, the crude strategy recommended RRT initiation more often in older patients with higher values of serum creatinine and serum potassium. On the second day, it seemed inclined to recommend RRT initiation in patients with stable arterial pH having a critical combination of low urine output and high blood urea nitrogen levels. Only on the third day did the learned strategies appear more aggressive recommending RRT initiation in most patients who had not recovered kidney function (i.e., patients with persisting low urine output and high blood urea nitrogen levels).

In this work, we chose to use HFD60 rather than mortality as the primary outcome. Mortality at a given timepoint conveys limited statistical information, as it contains only two possible values. In recent years, there has been an increased focus on patient-cantered non-mortality outcomes such as event-free day endpoints in ICU research (27, 32). In a dynamic reinforcement learning setting, there is however one more reason not to use survival as the outcome to optimize. Using survival as a distal reward signal may push the system to find a strategy that maximizes survival at the cost of unnecessary invasive procedures. Practically, the model could use its many degrees of freedom to learn a strategy that increases 60-day survival but decreases hospital and ICU discharge. On the contrary, optimizing over HFD60 is unlikely to yield longer hospital or ICU stays (33).

### Strength and limitations

To our knowledge, this study is the first to provide a validated dynamic decision support system for RRT initiation in the ICU. We believe the implications of our work are not only clinical but also methodological as the approach we used can be adapted for the timely initiation of a wide variety of treatments in medicine. However, our study has serval limitations. First, we considered only regular strategies, i.e., we did not allow for strategies to recommend stopping RRT before the third day if it had been initiated earlier. Disregarding the opportunities to stop treatment had a strong statistical advantage as it decreased the opportunities for a mismatch between prescribed and recommended treatments, thereby reducing variance in strategy learning and strategy evaluation. From a clinical standpoint, finding an optimal stopping strategy would rather be a distinct question that is more relevant after the third day. Second, we acknowledge that the effect size from implementing our learned strategies, though clinically relevant, was not statistically significant at the conventional 0.05 threshold. In reinforcement learning, learned strategies have long been tested on their training data, and inference for strategy evaluation is still rarely provided as reaching statistical significance often requires huge sample sizes (34). In this study, we performed external validation and estimated confidence intervals of the strategies’ benefits. This transparent approach indicates that developing more robust strategies may require training and testing on larger databases, perhaps coupling multiple electronic health records. Third, we concede that given infinitely large sample sizes, methods that leverage computation rather than expert knowledge (e.g., methods such as deep Q networks) may ultimately be more effective. Nevertheless, we believe that as even large electronic health records yield small effective sample sizes, encoding expert knowledge in the feature engineering process remains essential. Compared to black-box algorithms, we trust this human-centric approach is more likely to convince clinicians as it offers a window for interpretability.

### Implication for future research

As is true of traditional drugs, new individualized strategies will require proper testing in clinical settings before they can be deployed (35). This could be done for instance in a cluster randomized controlled trial comparing physicians alone to physicians assisted by the clinical decision support system. Alternatively, new trial designs could help to improve the learned strategy while it is being prospectively evaluated (36). Finally, if kidney damage markers (e.g., C-C motif chemokine ligand 14) demonstrate their clinical utility (37), new strategies leveraging this information may be developed. In the long run, these developments may help bridge the gap between biological knowledge and actionable data-driven approaches. We believe that fostering collaborations of clinical experts, methodologists, and mathematicians all genuinely interested in AKI and reinforcement learning is key. This, we hope, will continue to move personalized medicine forward for the benefit of intensive care patients.

In conclusion, we developed a dynamic RRT initiation strategy and confirmed via external validation that its implementation could increase the average number of days that ICU patients spend alive and outside the hospital. This interpretable strategy relies on routinely collected data and provides confidence intervals to guide decision-making at the bedside. It will require prospective testing and refinements before it can be broadly deployed in practice.

## Acknowledgements

The authors thank Cynthia T. Chen (Westaf) for editing. We thank all patients included in the AKIKI trials as well as their surrogates. We express our gratitude to the medical and nursing teams that participated in these trials.

## Authors’ contributions

FG, RP, FP, and VTT conceived the study. FG wrote the codes and did the computational analysis with input from RP and FP. SG, JPQ, and DD provided data from the AKIKI trials. ED designed the Sankey diagrams and contributed to the user interface. FG drafted the manuscript with inputs from RP, VTT, FP, SG, DD, and JPQ. All the authors read the paper and suggested edits. RP supervised the project. FG and RP accessed and verified the data. All authors had full access to all the data in the study and had final responsibility for the decision to submit for publication.

## ADDITIONAL INFORMATION

### Supplementary information

The online version contains supplementary material available at https://doi.org/XX.

**Supplementary Methods: Appendix A** Setup notations. **Appendix B** Summary of notations introduced in the appendix. **Appendix C** Doubly robust dynamic treatment regimen via weighted least squares. **Appendix D** Variable selection. **Appendix E** Missing data management. **Appendix E** Importance sampling for policy evaluation. **Appendix F** Advantage doubly robust estimator.

**Supplementary Results: Table S1.** Variance-covariance matrices of blip parameter estimates for the learned strategy based on multiple imputation analysis of one hundred data sets. **Figure S1.** Flow diagrams for the development set (A) and validation set (B). **Figure S2.** Comparison of recommendations from the original (A) or stringent (B) learned strategy and the RRT prescriptions received in the development set. **Figure S3.** Missing data patterns in the development set (A) and validation set (B).

#### Ethics approval and consent to participate

The MIMIC-III analysis received approval from the Institutional Review Boards of the Massachusetts Institute of Technology and Beth Israel Deaconess Medical Center (BIDMC). The AKIKI and AKIKI2 trials received approval from competent French legal authority (Comité de Protection des Personnes d’Ile de France VI, ID RCB 2013-A00765-40, NCT01932190 for AKIKI and ID RCB 2017-A02382-51, NCT03396757 for AKIKI 2) and consent of patient or relatives was obtained before inclusion.

#### Consent for publication

All authors have consented to the publication of the present manuscript, should the article be accepted by the Editor-in-chief upon completion of the refereeing process.

#### Availability of data and materials

The MIMIC-III data is publicly available at https://mimic.mit.edu. Anonymous participant data from the AKIKI trials is available under specific conditions. Proposals will be reviewed and approved by the sponsor, scientific committee, and staff on the basis of scientific merit and absence of competing interests. Once the proposal has been approved, data can be transferred through a secure online platform after the signing of a data access agreement and a confidentiality agreement.

## Supplementary Methods

### Setup notations

- *t*: decision timepoint, *t* ∈ {1,2,3}.
- *A_t_*: treatment observed at time *t*.
- *H_t_*: history of variables collected up to time *t* including the treatment received before time *t* but excluding the treatment received at time *t*.
- 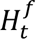: subset of *H_t_* relevant for prognosis.
- 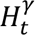: subset of *H_t_* relevant for the effect of treatment initiation at time *t*.
- *Y*: outcome of interest.
- 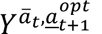: potential outcome corresponding to the outcome that would have been observed if treatments *a*_1_,…,*a_t_* had been delivered at decision timepoints 1,…,*t* and (possibly contrary to fact) all subsequent treatment decisions had been optimal. We use over and underline notations to indicate the past and future treatments respectively i.e., 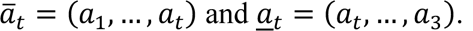.

### Summary of notations introduced in the appendix

- 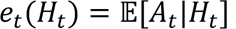: propensity score at time *t*.
- 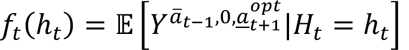: treatment-free function at time *t*.
- 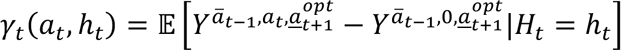: blip function at time *t*.
- 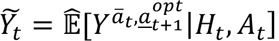: pseudo-outcomes at time *t*.
- 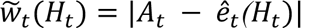: overlap weights at time *t*.
- τ = (*H*_3_, *A*_3_, *Y*): the observable full trajectory of a patients.
- *T*: the space of trajectories.
- 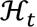: the space of histories of variables collected up to time *t*.
- π = (π_1_, π_2_, π_3_) with 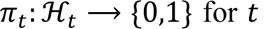 for *t* = 1,2,3: the non-stationary deterministic policy*π.
- 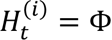: indicates that at time *t*, patient *i* is in a terminal state (i.e., death).

### Doubly robust dynamic treatment regimen via weighted least squares

The procedure (termed dWOLS) was formally introduced and described in detail by Wallace and Moodie.^1^ Succinctly, the method requires that for each decision timepoint *t* = 1,2,3, we posit models for the propensity scores 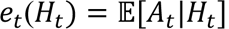 as well as the treatment-free *f_t_*(·), and blip γ*_t_*(·) functions. Treatment-free and blip functions are defined as 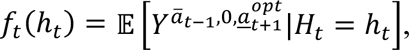 and 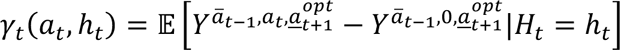 so that 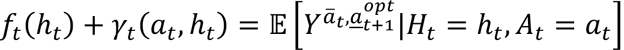.^†^ The estimation of *f_t_*(·) and γ*_t_*(·) starts at *t* = 3 by regressing 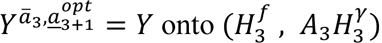 via weighted least squares with weights 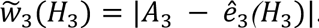. The procedure then follows a backward stepwise approach where we substitute all unobserved potential outcomes by pseudo-outcomes. Specifically, for *t* = 2,1, we build pseudo-outcomes 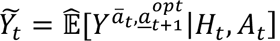 by taking naive outcomes *Y* and summing up subsequent regrets i.e., *Y*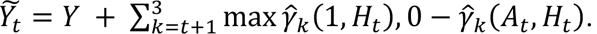. Pseudo-outcomes at time *t* represent the outcomes that would have been observed if treatment decisions had been optimal from time *t* + 1 onwards. We then regress *Y*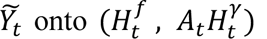 via weighted least squares with weights 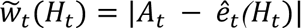. Using these overlap weights provide double robustness and enhance sample efficiency. Note that the dWOLS estimation procedure does not require making a Markovian assumption. It only requires assuming that for each decision timepoints either the variables causing renal replacement therapy (RRT) initiation, or the prognosis variables were measured. Because we considered that once initiated, RRT is not stopped in the three days following stage 3 KDIGO-AKI, analysis for each decision timepoint was limited to those participants who had not initiated RRT until this decision timepoint (as those who had initiated RRT had no treatment decision to make).

### Variable selection

For each decision timepoint, the variables we considered for modeling the probability of RRT initiation withing 24 hours were: blood urea nitrogen, serum potassium, arterial blood pH, and urine output. For each decision timepoint, the variables we considered for predicting hospital-free days at day 60 (HFD60) were: age, weight, gender, SOFA score, serum creatinine, blood urea nitrogen, serum potassium, arterial blood pH, and urine output. We considered the evolving values of the aforementioned variables prior to the decision timepoint of interest. The same variables were considered in development and validation analyzes.

### Missing data management

Missing data were handled through multiple imputations by chained equations using outcomes as well as all aforementioned predictors in the imputation models. One hundred independent imputed data sets were generated and analyzed separately. Variance-covariance matrices of blip functions parameters were estimated using the bootstrap (999 iterations). Estimates were then pooled using Rubin’s rules.

### Importance sampling for policy evaluation

We used importance sampling for policy evaluation in reinforcement learning^2^ to estimate hospital mortality as well as the proportion of patients who would initiate RRT within three days under a learned strategy. The approach is similar to inverse propensity weighting as used in the context of marginal structural modeling in epidemiology.^3^ Succinctly, denoting τ = (*H*_3_, *A*_3_, *Y*) ∈ *T* the observable full trajectory of a patient and 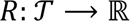 any reward function of the trajectory, the expected reward under a different strategy, say the non-stationary deterministic strategy π i.e., π = (π_1_, π_2_, π_3_) with 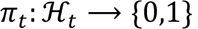 for *t* = 1,2,3, can be estimated by

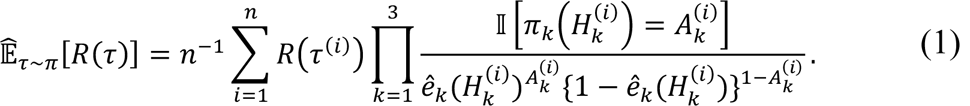

To estimate hospital mortality, the reward function we used was *R*(τ^(*i*)^) = 1 if patient *i* died in the hospital and *R*(τ^(*i*)^) = 0 otherwise. To estimate the proportion of patients who would initiate RRT within three days under our learned strategies, we used the reward function 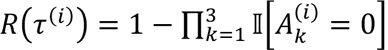: which outputs one whenever patient *i* initiated RRT at any time along their observed trajectory. The estimator above straightforwardly handles the patients who died before day 3, provided we consider that the strategy π stops prescribing treatment once a patient has died i.e., π*_k_*(Φ) = 0, and that no RRT was prescribed to the patients who have died i.e., *e_k_*(Φ) = 0.^‡^ The variables we considered for modeling the propensity scores are identical to those given in the previous section. To improve efficiency, we used the weighted version of the estimator above that is given in equation 3 from Precup et al.^2^

### Advantage doubly robust estimator

Although the estimator given in equation 1 accounts for the patients who died before day 3, it only uses trajectories that match the policy π exactly, which can make policy evaluation sample inefficient. To estimate the causal effect of implementing the original, stringent, or treat all strategies compared to current best practices, we used the cross-fitted advantage doubly robust (ADR) estimator with terminal state for strategy evaluation given in the Algorithm 2 from Nie et al.^4^ The ADR estimator allows the evaluation of when-to-treat-policies exploiting the subparts of trajectories that match the policy π. The original, stringent, and treat all strategies are all regular when-to-treat policies in the sense of Definition 1b from Nie et al. The ADR estimator is more data efficient but also more robust than the estimator given in equation (1). Briefly, this estimator relies on the decomposition into a sum of local advantages of the relative value of any given strategy in comparison to that of the never-treating policy **0**, following Lemma 1 of Murphy.^5^ The ADR estimand is 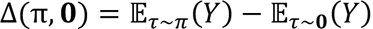 where, π denotes the strategy to be tested, and zero is the never-treating policy. The causal effects of implementing the original, stringent, or treat all strategies compared to a “best practices policy” denoted π_*bp*_, are given by

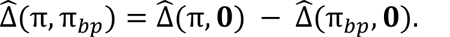

As in the dWOLS procedure, the ADR estimator does not need any structural (e.g., Markovian) assumptions. As Nie et al.,^4^ we estimated all the nuisance components using cross-fitting to reduce the effect of own-observation bias.

For each decision timepoint, the variables we considered for modeling the probability of RRT initiation withing 24 hours were: blood urea nitrogen, serum potassium, arterial blood pH, and urine output. For each decision timepoint the variables we considered for predicting hospital-free days at day 60 (HFD60) were: age, weight, gender, SOFA score, serum creatinine, blood urea nitrogen, serum potassium, arterial blood pH, and urine output.

Missing data were handled through multiple imputations by chained equations using outcomes as well as all aforementioned predictors in the imputation models. Twenty independent imputed data sets were generated and analyzed separately. The variances of the estimators were estimated using the bootstrap (999 iterations). Estimates were then pooled using Rubin’s rules.

## Supplementary Results

**Table S1.**
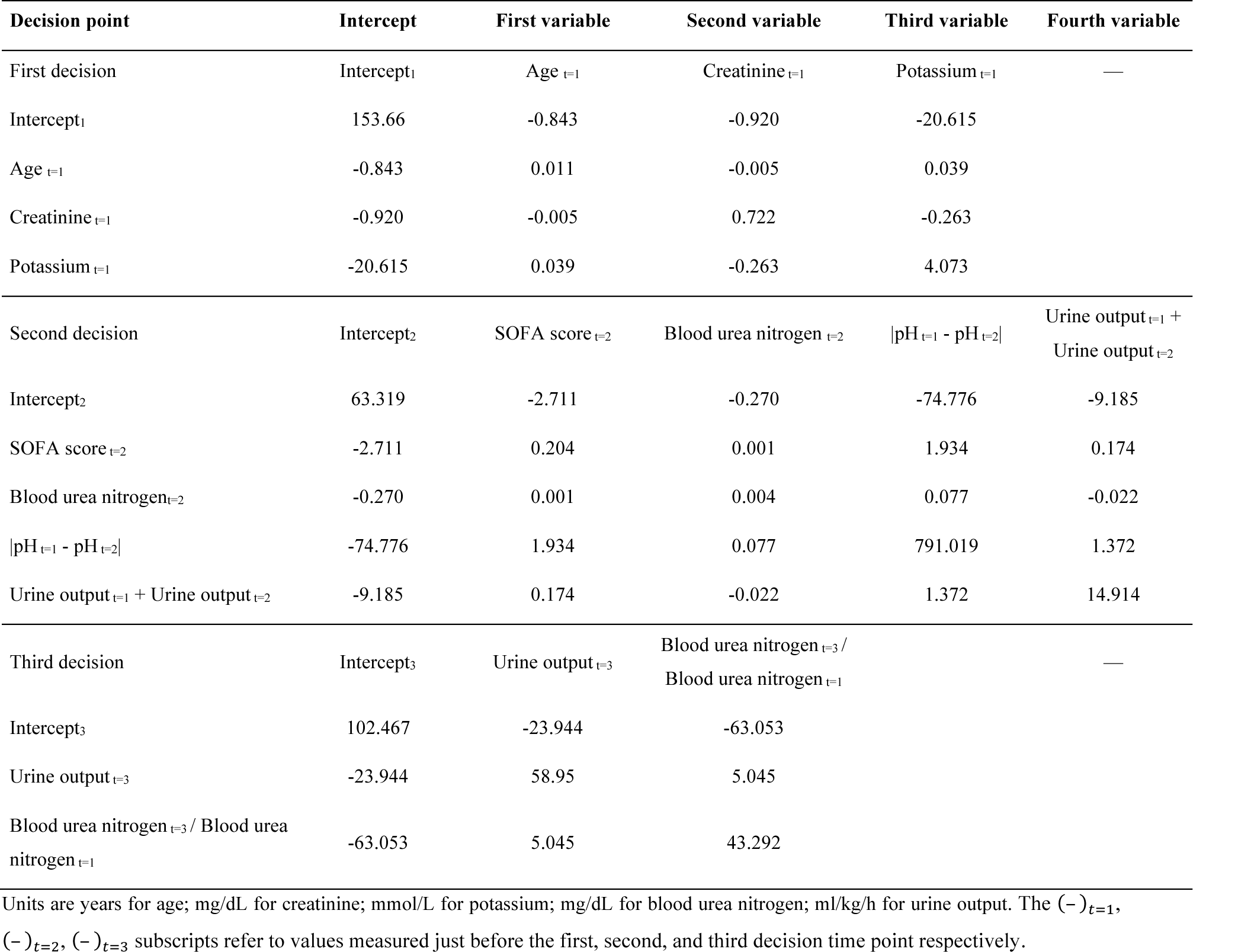
Variance-covariance matrices of blip parameter estimates for the learned strategy based on multiple imputation analysis of one hundred data sets. Denoting *H_t_* a patient’s vector of covariates at decision timepoint 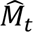 the estimated variance-covariance matrix from decision timepoint 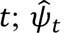 the blip parameter estimates from decision timepoint *t*, 95% confidence intervals for the individual blips can be calculated as

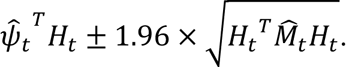

**Figure S1.**
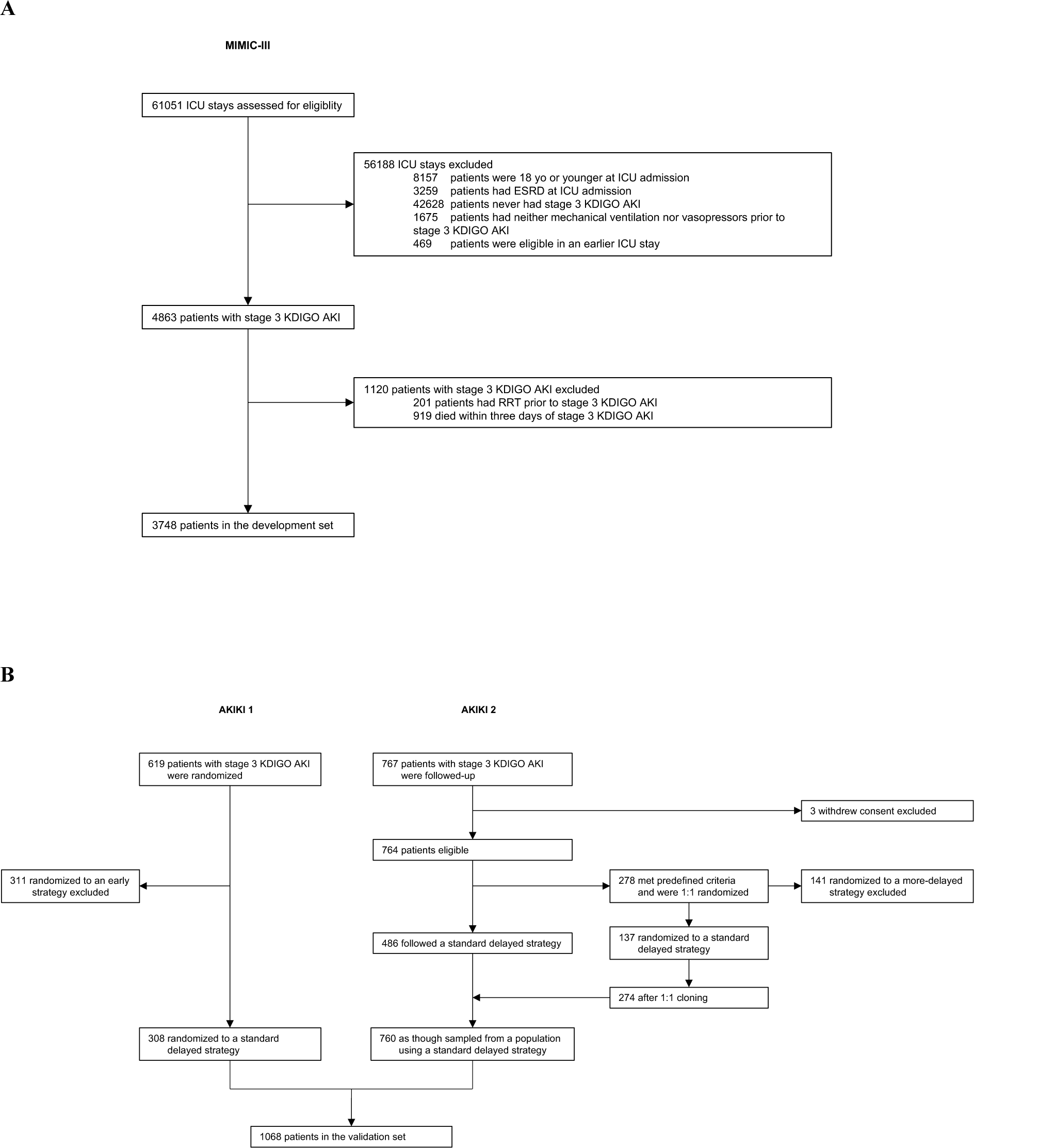
Flow diagrams for the development set (A) and validation set (B).

**Figure S2.**
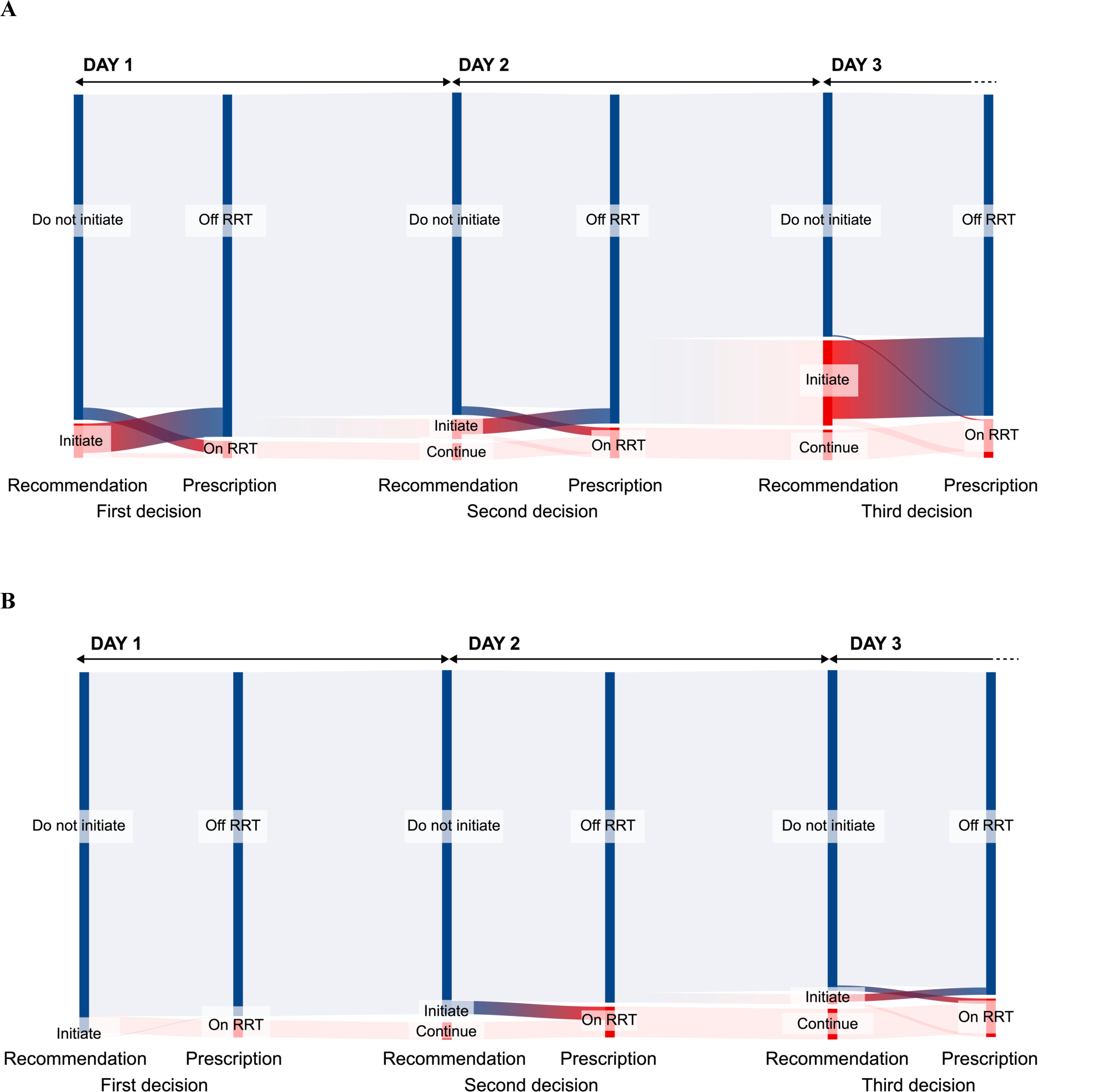
Comparison of recommendations from the original (A) or stringent (B) learned strategy and the RRT prescriptions received in the development set.

**Figure S3.**
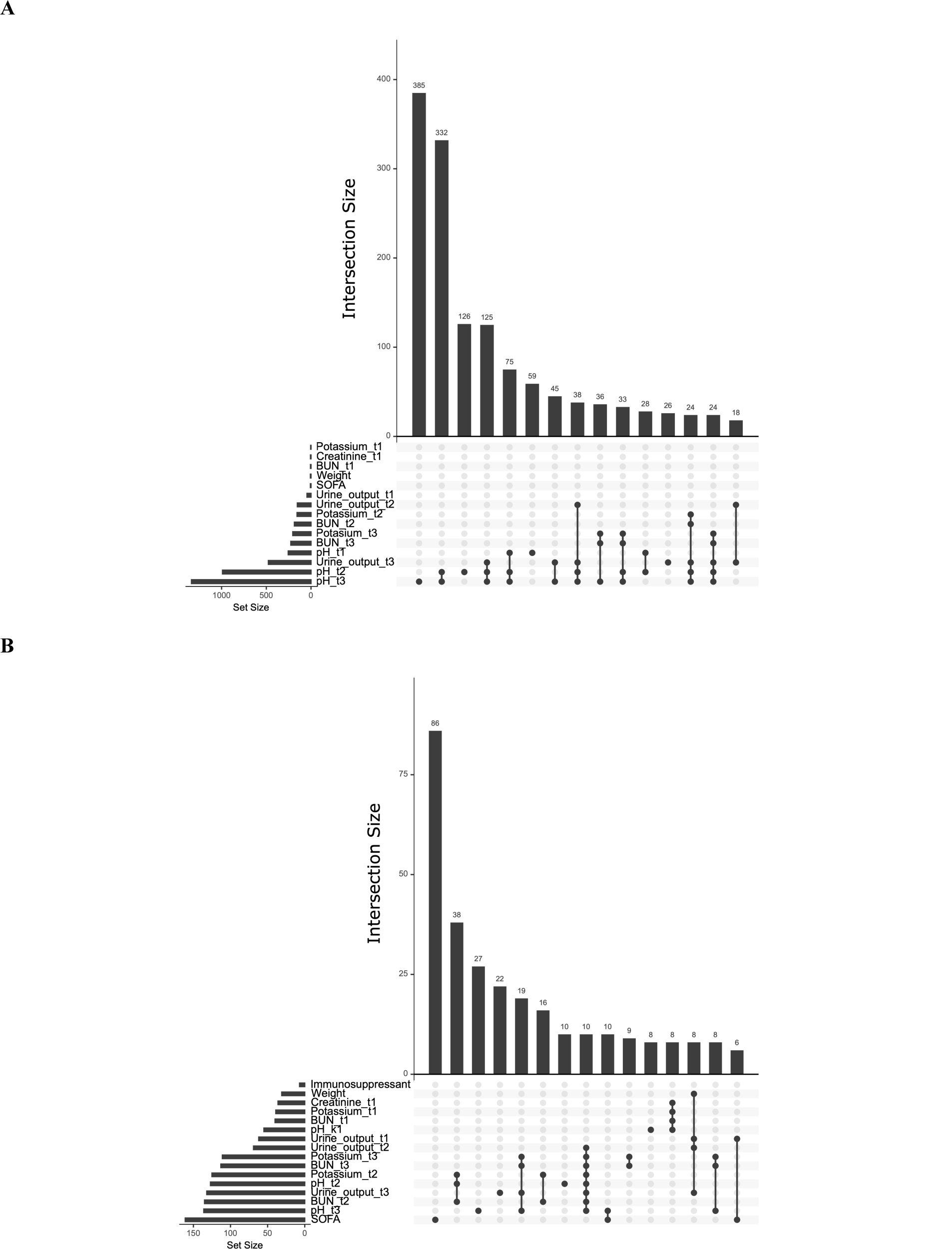
Missing data patterns in the development set (A) and validation set (B).

## ABREVIATIONS

AKI: Acute Kidney Injury
BIDMC: Beth Israel Deaconess Medical Center CI: Confidence Interval
HFD60: Hospital-Free Days at day 60 ICU: Intensive Care Unit
IQR: Interquartile Range
KDIGO: Kidney Disease: Improving Global Outcomes RCT: Randomized Controlled Trial
RRT: Renal Replacement Therapy
SMART: Sequential Multiple Assignment Randomized Trial SOFA: Sequential Organ Failure Assessment

## Footnotes

* Note that throughout the paper, we use the term strategy rather than policy. For the remainder of this appendix, these can be taken to be synonymous.

† This last equality uses the sequential ignorability assumption i.e., 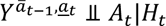 for all *t*.

‡ For the sake of clarity, we denoted 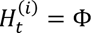 when patient *i* is in a terminal state (i.e., death) at time *t*.

